# Higher serological responses and increased vaccine effectiveness demonstrate the value of extended vaccine schedules in combatting COVID-19 in England

**DOI:** 10.1101/2021.07.26.21261140

**Authors:** Gayatri Amirthalingam, Jamie Lopez Bernal, Nick J Andrews, Heather Whitaker, Charlotte Gower, Julia Stowe, Elise Tessier, Vani Subbarao, Georgina Ireland, Frances Baawuah, Ezra Linley, Lenesha Warrener, Michelle O’Brien, Corinne Whillock, Paul Moss, Shamez N Ladhani, Kevin E Brown, Mary E Ramsay

**Affiliations:** Immunisation and Countermeasures Division, National Infection Service, Public Health England, London, United Kingdom; Statistics, Modelling and Economics Department, National Infection Service, Public Health England, London, United Kingdom; Brondesbury Medical Centre, Kilburn, London, United Kingdom; Sero-Epidemiolgy Unit, National Infection Service, Public Health England, Manchester, United Kingdom; Virus Reference Department, National Infection Service, Public Health England, London, United Kingdom; Institute of Immunology and Immunotherapy, University of Birmingham, Edgbaston, United Kingdom; Paediatric Infectious Diseases Research Group, St. George’s University of London, London, United Kingdom

**Keywords:** COVID-19, COVID-Vaccine, Antibody, Spike Protein, Immunity, Pfizer, AstraZeneca

## Abstract

**Introduction:** In January 2021, the UK decided to prioritise the delivery of the first dose of BNT162b2 (Pfizer/BioNTech) and AZD1222 (AstraZeneca) vaccines by extending the interval until the second dose up to 12 weeks.

**Methods:** Serological responses were compared after BNT162b2 and AZD1222 vaccination with varying intervals in uninfected and previously-infected adults aged 50-89 years. These findings are evaluated against real-world national vaccine effectiveness (VE) estimates against COVID-19 in England.

**Results:** We recruited 750 participants aged 50-89 years, including 126 (16.8%) with evidence of previous infection; 421 received BNT162b2 and 329 and AZD1222. For both vaccines, over 95% had seroconverted 35-55 days after dose one, and 100% seroconverted 7+ days after dose 2. Following a 65-84 day interval between two doses, geometric mean titres (GMTs) at 14-34 days were 6-fold higher for BNT162b2 (6703; 95%CI, 5887-7633) than AZD1222 (1093; 806-1483), which in turn were higher than those receiving BNT162b2 19-29 days apart (694; 540 - 893). For both vaccines, VE was higher across all age-groups from 14 days after dose two compared to one dose, but the magnitude varied with interval between doses. Higher two-dose VE was observed with >6 week intervals between BNT162b2 doses compared to the authorised 3-week schedule, including ≥80 year-olds.

**Conclusion:** Our findings support the UK approach of prioritising the first dose of COVID-19 vaccines, with evidence of higher protection following extended schedules. Given global vaccine constraints, these results are relevant to policymakers, especially with highly transmissible variants and rising incidence in many countries.

**Funding:** Public Health England

## Background

Older adults have been disproportionately affected by the COVID-19 pandemic, with age being the single most important risk factor for hospitalisations and deaths.(1-3) In the United Kingdom, older adults were prioritised for vaccination at the start of the COVID-19 immunisation programme on 08 December 2020, initially with the Pfizer/BioNTech (BNT162b2) vaccine using the authorised 3-week interval between doses.(4) From 04/01/2021, the AstraZeneca (AZ) vaccine (AZD1222) was deployed and, with its more favourable storage and transport conditions, was used for vaccinating in care homes, community healthcare professionals and healthy adults aged 40-60 years. In January 2021, the UK Joint Committee on Vaccination and Immunisation (JCVI) recommended that, with the emergence of a second wave, the second dose of vaccine should be extended for up to 12 weeks to prioritise the first dose for those at highest risk of severe COVID-19 and death.(5) The decision to extend the second dose was based on early clinical trial data indicating nearly 90% effectiveness against SARS-CoV-2 within 3 weeks of the first dose of BNT162b2 vaccine compared to 95% from two weeks after the second dose.(6) Vaccinating more at-risk individuals quickly with a single dose was predicted to prevent more cases, hospitalisations and deaths than two doses at a 3-week interval.(7) This unique approach against authorised use and without formal clinical trials resulted in considerable international debate and prompted the need to evaluate immune responses and vaccine effectiveness following extended schedules.

The COVID-19 vaccine responses after extended immunisation schedules (CONSENUS) evaluation aimed to assess immune responses in ≥50 year-olds receiving a COVID-19 vaccine as part of the UK extended immunisation schedule. Early analysis indicated that a single dose of BNT162b2 vaccine was associated with >94% seropositivity after 3 weeks in previously uninfected older adults, while two doses produced very high antibody levels, significantly higher than convalescent sera from adults with mild-to-moderate PCR-confirmed COVID-19.(5) Real world effectiveness studies indicate 50-70% protection against infection or mild disease for ≥8 weeks after one BNT162b2 dose and ≥6 weeks after AZD1222, with 75-85% protection against hospitalisation or death.(8)

We now report serological responses in 750 adults aged 50-89 years given two doses of BNT162b2 or AZD1222 at different intervals, comparing serological responses. These findings are evaluated against real-world vaccine effectiveness estimates against COVID-19 using similar dosing intervals in the same age-group in England.

## Method

### Participants

CONSENSUS recruited immunocompetent adults aged ≥50 years in January 2021 through London primary care networks to provide serial blood samples at 0,3,6,9,12,15 and 20 weeks after their first dose of COVID-19 vaccine. As part of the national COVID-19 vaccine roll out, participants received either (i) two BNT162b2 doses at 3-4 weeks apart (ii) two BNT162b2 doses up to 12 weeks apart or (iii) two AZD1222 doses up to 12 weeks apart. Antibody responses were compared with convalescent samples from adults with mild-to-moderate PCR-confirmed COVID-19, up to 98 days after symptom onset.

### Serological Testing

Serum samples were tested for nucleoprotein (N) antibodies as a marker of previous SARS-CoV-2 infection (Anti-SARS-CoV-2 total antibody assay, Roche Diagnostics, Basel, Switzerland) and spike (S) protein antibodies, which could be infection- or vaccine-derived (Elecsys Anti-SARS-CoV-2 S total antibody assay, Roche Diagnostics: positive ≥ 0.8 arbitrary units (au)/mL to assess vaccine response).(9, 10)

### Assessment of Vaccine Effectiveness

A test negative case-control design was used to estimate odds ratios for testing SARS-CoV-2 positive to in vaccinated compared with unvaccinated people with COVID-19 compatible symptoms who were tested using polymerase chain reaction (PCR), as described previously.(8)

### Data sources

#### Outcome assessment

All healthy adults aged ≥50 years in England were eligible for inclusion. Testing for COVID-19 in the UK is done through hospital and public health laboratories (pillar 1) and more widely through community (pillar 2) testing.(11) Pillar 2 tests performed between 26/10/2020 and 18/06/2021 we extracted for those who reported being symptomatic.

#### Exposure assessment

Testing data were linked to individual vaccination histories in the National Immunisation Management System (NIMS), using unique National Health Service numbers, date of birth, surname, first name, and postcode. NIMS data were extracted on 21/06/2021 with immunisation records up to 20/06/2021.(12)

### Statistical Analysis

#### Serological Assessment

Geometric mean antibody titres (GMTs) were calculated with 95% confidence intervals (CI). Geometric mean ratios (GMR) of responses between timepoints were estimated using a regression model on log responses including a random effect for each participant. The GMR of responses by vaccine type at each post-vaccination timepoint was estimated via regression on log Roche S responses and included age-group and sex. Statistical analyses were performed using STATA v.14.2. Individuals testing positive on the Roche N assay were considered to have had prior SARS-CoV-2 infection. Infection status was changed if a seronegative participant seroconverted on the Roche N assay during the study and remained positive thereafter.

#### Vaccine Effectiveness

Logistic regression was used to estimate the odds of vaccination in PCR confirmed cases compared with those who tested negative for SARS-CoV-2. Only those swabbed within 0-10 days of symptom onset were included in the analysis because sensitivity of PCR testing decreases beyond 10 days after symptom onset. Vaccine effectiveness was calculated as 1 minus odds ratio.

To estimate vaccine effectiveness in fully susceptible people, we excluded those with a previous positive PCR or antibody result prior to 8 December 2020. As previously described (8), estimates were adjusted for week of onset, 5-year age bands, gender, NHS region, index of multiple deprivation (quintiles), ethnicity, health/social care worker, care home resident, flagged as clinically extremely vulnerable in NIMS, and flagged as being in the extended risk-groups in NIMS among <65 year-olds only. Analyses were run separately for 50-64, 65-79, 80+ and 80+ (early cohort) year-olds. For 50-64 year-olds, only unvaccinated or vaccinated individuals from 01/02/2021 were included because, before this, only health and social care workers were eligible in this age-group. For age 65-79 and 80+ year-olds, a cut-off date of 04/01/2021 was used when AZD1222vaccine became available. The 80+ early-cohort age-group included only those unvaccinated or first dose vaccinated before 04/01/2021 with the BNT162b2 vaccine, mostly with a 3-week interval between doses.

VE was estimated by vaccine manufacturer and according to intervals after first dose as well as from 14 days after the second dose split by intervals between first and second doses of 19-29,30-44,45-64,65-84 and 85+ days.

### Role of funding source

The study was self-funded by Public Health England.

### Ethics Approval

The protocol was approved by PHE Research Ethics Governance Group (reference NR0253; 18/01/2021).

## Results

### Participants

We recruited 750 participants aged 50-89 years (median age, 71, IQR 66-76 years)- 421 received at least one BNT162b2 dose and 329 at least one AZD1222 dose (Table 1). Overall, 46% (344/746) were male, 27% (171/743) were of non-White ethnicity, 16.8% (126/750) had evidence of previous infection at enrollment and one seroconverted during the study. Adults aged 50-64 years were more likely to have evidence of previous infection than older adults (56/171; 32.8% vs 70/579; 12.1%; p<0.001).

**Table 1:**
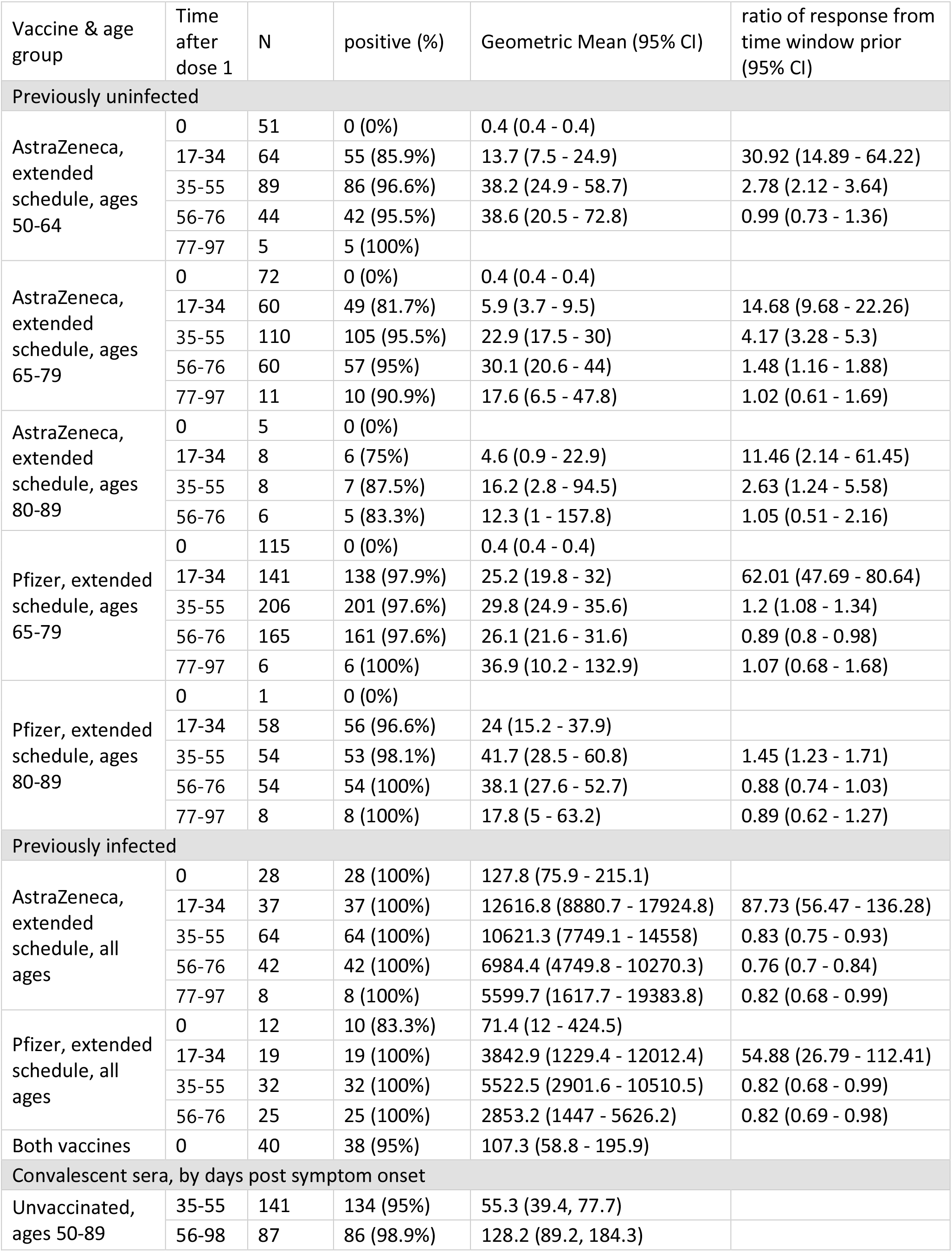
Proportion Roche S seropositive, Geometric Mean Concentrations (if >5 samples) by vaccine type and age group and ratio of responses by time, following dose 1

### Post dose 1: Antibody responses in uninfected adults

Among BNT162b2 vaccine recipients receiving an extended schedule, seropositivity increased rapidly after the first dose, with 97.7% (217/222) seroconverting by 17-34 days and 35-55 days (97.7%;254/260) post-vaccination. S-antibody GMTs peaked by 35-55 days after vaccination at 29.8 (95%CI: 24.9-35.6) for 64-79 year-olds and 41.7 (95%CI: 28.5-60.8) for 80-89 year-olds, with levels sustained to 77-97 days post-dose 1 (Figure 1, Table 2).

**Table 2:** Geometric mean ratio of responses, adjusted for age and sex, following dose 1 of an extended vaccine schedule

|  | 0 weeks |  | 17-34 days |  | 35-55 days |  | 56-76 days |  |
| --- | --- | --- | --- | --- | --- | --- | --- | --- |
|  | geometric mean ratio of (log) RocheS responses | p-value | geometric mean ratio of (log) RocheS responses | p-value | geometric mean ratio of (log) RocheS responses | p-value | geometric mean ratio of (log) RocheS responses | p-value |
| vaccine |  |  |  |  |  |  |  |  |
| Astra Zeneca | 1(ref) |  | 1(ref) |  | 1(ref) |  | 1(ref) |  |
| Pfizer | 0.99<br>(0.98 - 1) | 0.171 | 4.05<br>(2.49 - 6.6) | <0.001 | 1.39<br>(1 - 1.93) | 0.049 | 1<br>(0.68 - 1.48) | 0.987 |
| age group |  |  |  |  |  |  |  |  |
| 50-64 | 1(ref) |  | 1(ref) |  | 1(ref) |  | 1(ref) |  |
| 65-79 | 1<br>(0.99 - 1.01) | 0.928 | 0.49<br>(0.27 - 0.9) | 0.021 | 0.58<br>(0.38 - 0.87) | 0.009 | 0.68<br>(0.4 - 1.15) | 0.151 |
| 80-89 | 0.99<br>(0.97 - 1.02) | 0.692 | 0.47<br>(0.22 - 0.98) | 0.045 | 0.69<br>(0.39 - 1.21) | 0.196 | 0.8<br>(0.42 - 1.52) | 0.492 |
| sex |  |  |  |  |  |  |  |  |
| male | 1(ref) |  | 1(ref) |  | 1(ref) |  | 1(ref) |  |
| female | 1<br>(0.99 - 1.01) | 0.481 | 1.24<br>(0.84 - 1.82) | 0.273 | 1.85<br>(1.41 - 2.43) | <0.001 | 1.72<br>(1.26 - 2.34) | 0.001 |

**Figure 1:**
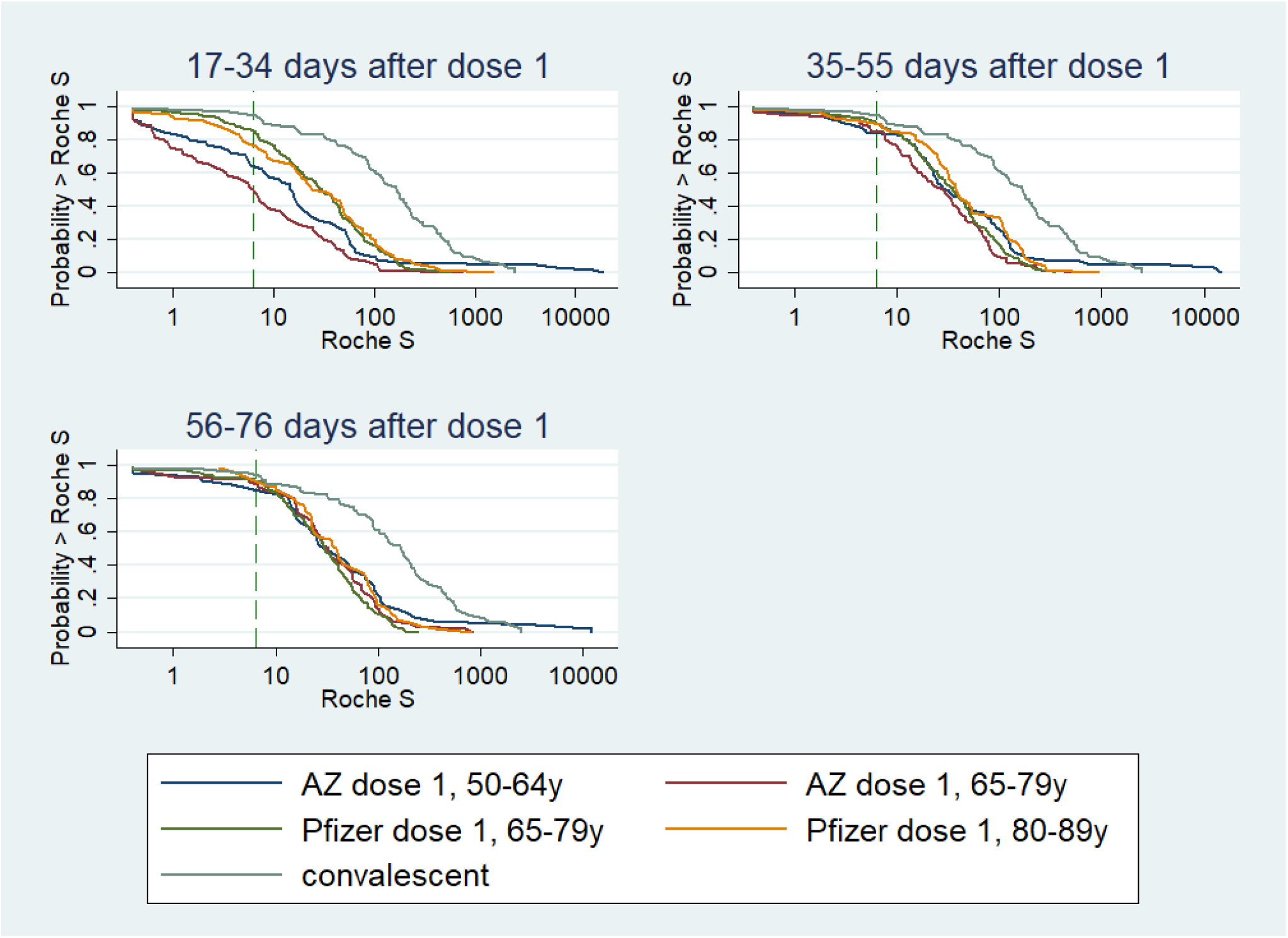
Reverse Cumulative Distribution curves, antibody responses following 1st dose of COVID-19 vaccine in previously uninfected individuals, by vaccine, age group, and including a curve for unvaccinated convalescent cases 56-98 days post-infection

Among AZD1222 vaccine recipients, 85.9% (55/64) of 50-64 year-olds and 81.7% (49/60) of 65-79 year-olds seroconverted at 17-24 days, which increased to 95.6% (198/207) overall at 35-55 days. GMTs continued to increase from 13.7 (95%CI: 7.5-24.9) at 17-34 days to 38.2 (95%CI: 24.9-58.7) at 35-55 days in 50-64 year-olds, whilst, in older adults the GMT peak was delayed until 56-76 days. The GMR for S-antibody was 62.01 (95%CI: 47.69-80.64) for BNT162b2 vaccine recipients aged 65-79 years at 17-34 days compared to pre-vaccine, followed by a GMR of 1.2 (95%CI: 1.08-1.34) from 17-34 days to 35-55 days after dose 1. For the AZD1222 vaccine in the same age-group, GMRs were 14.68 (95%CI: 9.68-22.26) and 4.17 (95%CI: 3.28-5.3), respectively. In previously-uninfected individuals, GMTs remained lower in the 13 weeks post-dose 1 for both vaccines compared to convalescent sera from mild-to-moderate PCR-confirmed cases (Figure 1, Table 1).

GMRs between BNT162b2:AZD1222 in previously-uninfected recipients was 4.05 (95%CI: 2.49-6.6) at 17-34 days after dose 1, this ratio declined with time and was no longer statistically significant at 56-76 days post-vaccination (Table 2, Figure 1). Females had higher S-antibody GMTs than males, while differences in age-groups narrowed with time since vaccination such that there was no significant difference by age-group at 56-76 days post-vaccination (Table 2, Figure 1).

### Post dose 1: Vaccinees with previous COVID-19 infection

In adults with serological evidence of prior infection, S-antibody levels at vaccination were not significantly different from convalescent sera at 56-98 days post-infection (Table 2). S-antibody GMTs increased from 71.4 (95%CI: 12.0-424.5) to 3,842.9 (95%CI: 1229.4-12012.4) at 17-34 days post-vaccination for BNT162b2 recipients, and from 127.8 (95%CI: 75.9-215.1) to 12616.8 (95%CI: 8880.7-17924.8) for AZD1222 vaccine recipients. These initially high titres subsequently waned through 56-76 days after dose 1.

### Post dose 1: Vaccine effectiveness sustained ≥8 weeks following dose 1

The odds of testing SARS-CoV-2 PCR-positive among vaccinated people increased up to days 7-9 after dose 1 for both vaccines, reaching 1.12 (VE: -12%) and 1.19 (VE: -19%) for AZD1222 and BNT162b2 in 65-79 year-olds, respectively (Figure 2). Among ≥80 year-olds receiving BNT162b2, VE increased from days 14-20, reaching 61% (95%CI: 49-71) in the early cohort (three-week interval) and 52% (95%CI: 39-63) in the later (longer interval) cohort at 28-34 days and remained at similar levels between days 35-55 (5-8 weeks). Amongst 65-79 year-olds, VE began to increase from 10-13 days after vaccination, reaching 53% (95%CI: 45-60) on days 28-34, and remained at a similar level between 35-69 days (5-10 weeks). A similar trend was observed in the BNT162b2 recipients aged 50-64 years with a VE of 58% at days 28-34. Whilst there was some evidence of a 10-20% decrease in VE by 10 weeks after the first dose, there was an apparent rise again in VE at the final interval, although with wide confidence intervals (Figure 2).

**Figure 2:**
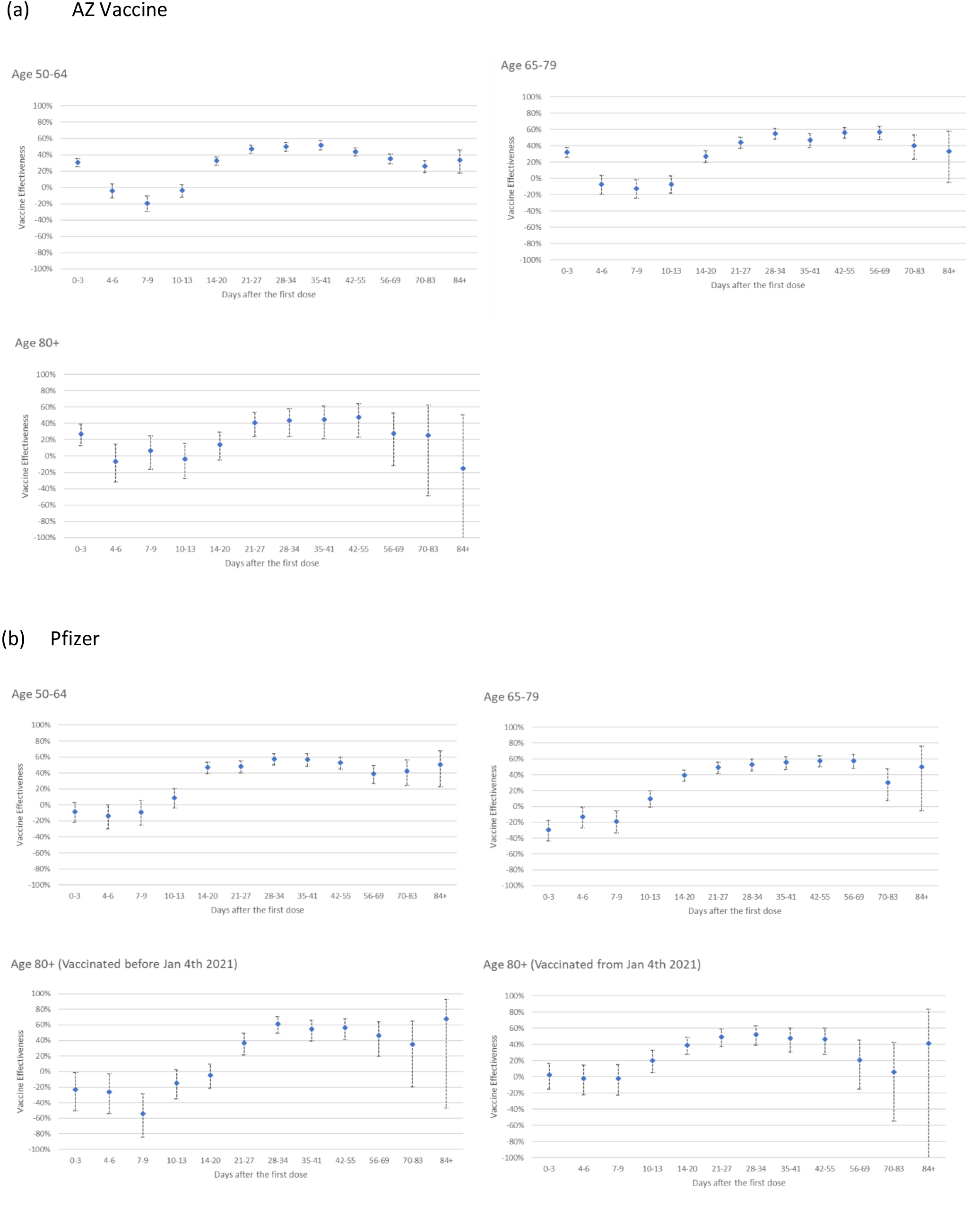
Adjusted vaccine effectiveness against confirmed symptomatic COVID-19 by interval after vaccination amongst 50-64 year olds, 65-79 year olds and 80+ year olds with the (a) AZ vaccine and (b) Pfizer-BioNTech BNT162b2 vaccine

For AZD1222 vaccine, the positive VE within 3 days of vaccination was likely an artefact because vaccinated adults were getting PCR-tested and reported test-negative due to vaccine reactogenicity (Figure 2). In adults aged ≥80 years, VE increased from days 14-20, reaching 43% (95%CI: 24-58) on days 28-34 and remained at a similar level between days 35-55 (5-8 weeks). Amongst 65-79 year-olds, VE increased from days 14-20 post-vaccination, reaching 55% (95%CI: 48-61) after 28 days, and then remaining stable until days 56-69 after the first vaccine dose. A similar trend was observed among 50-64 year-olds, with 50% (95%CI: 45-55) VE estimates. A reduction in VE was noted by 70 days post-vaccination at 40% (95%CI: 23-53) and 26% (95%CI: 18-33) for 65-79 year-olds and 50-64 year-olds, respectively. For ≥80 year-olds, confidence intervals were too wide to asses declines, but point estimates showed a decline after 8 weeks.

### Post dose 2: Antibody responses by dosing Interval

Vaccine dose intervals varied between 3-13 weeks amongst CONSENSUS participants. They were, therefore, assessed by the following intervals: (i) 19-29 (ii) 45-64 (iii) 65-84 and (iv) ≥85 days. Sampling timepoints post-dose 2 were divided into 7-13 and 14-34 days. The vaccine used and dose intervals varied by age-group depending on national recommendations and vaccine supply. Those receiving BNT162b2 at 19-29 day intervals and those reaching 85+ days for either vaccine were older (median age 76, 76 and 74 years, respectively) whilst those receiving AZD1222 at 45-65 and 65-84 intervals were younger (66 and 66.5 years, respectively) because initial vaccine rollout prioritised older adults and recommended a 3-week interval between doses.

Regardless of vaccine and schedule, all participants seroconverted 7+ days after dose 2 (AZD1222: N=200; BNT162b2 extended: N=282 and BNT162b2 control: N=87). BNT162b2 dose 2 responses were quick, peaking at 7-13 days followed by a 23% decline at 14-34 days. Amongst BNT162b2 recipients, GMTs were 10-fold higher at 14-34 days post-dose 2 following a 65-84 day interval compared with a 19-29 day interval (Table 3, Figure 3). Furthermore, among those with a vaccine interval of 85+ days, GMTs at 7-13 days post-dose 2 were higher compared with 65-84 days. There were, however, too few results to confirm this trend beyond 14 days post-dose 2.

**Table 3:**
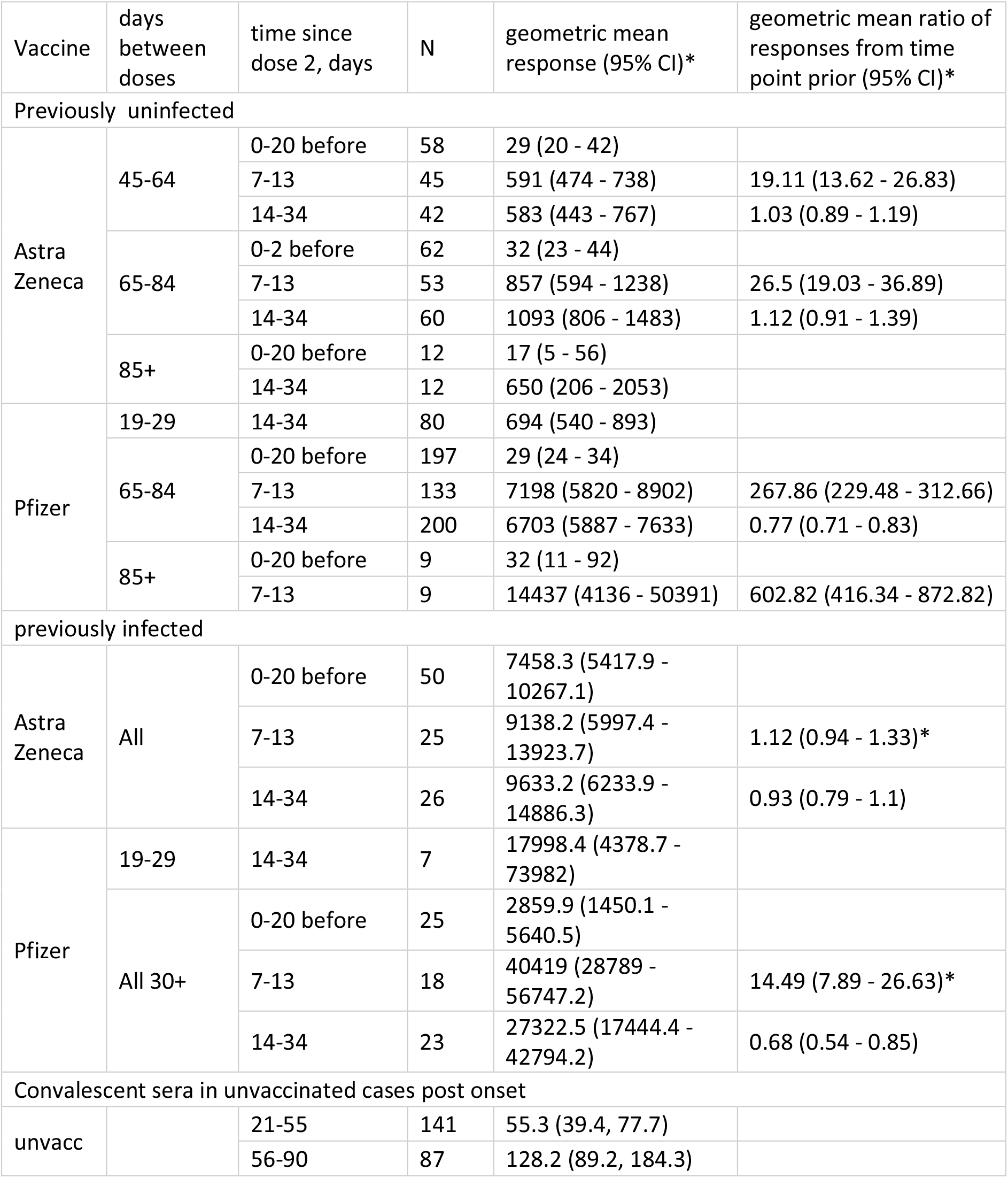
Geometric mean responses and geometric mean ratio of responses following dose 2, by vaccine, interval compared with convalescent sera *\*GMRs at 7-13 days post two are relative to responses at 0-20 days before dose 2*

**Figure 3:**
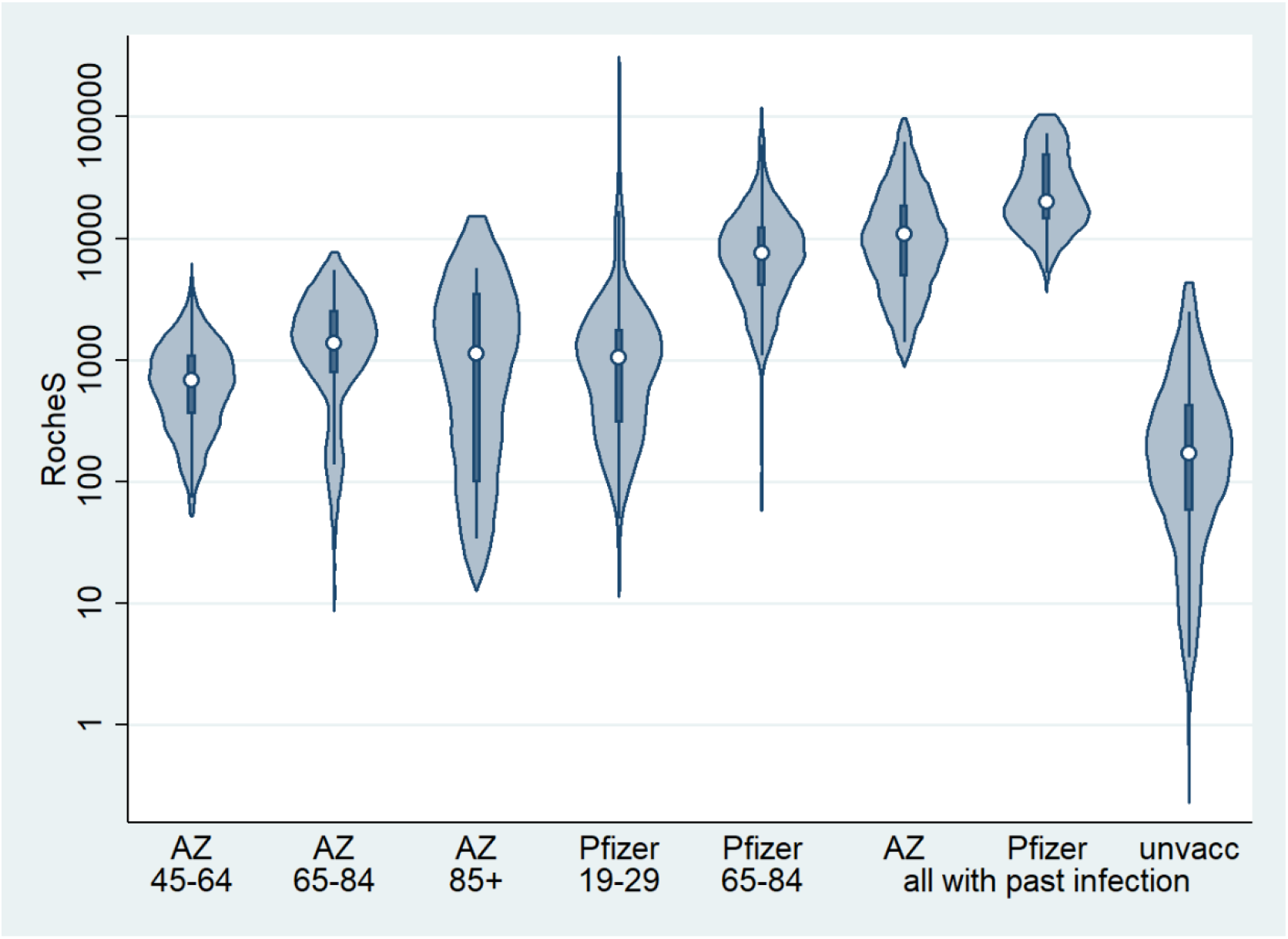
Violin plots, antibody responses at 14-34 days following 2^nd^ dose of COVID-19 vaccine in (i) previously uninfected individuals, by vaccine and interval between doses, (ii) in previously infected individuals by vaccine (any schedule) and (iii) in unvaccinated convalescent individuals.

GMTs with a 65-84 day interval were 6-fold higher after BNT162b2 (6703; 95%CI, 5887–7633) compared to AZD1222 (1093; 95%CI: 806-1483) at 14-34 days post-dose 2 (Table 4). However, GMTs among AZD1222 recipients with an extended schedule were significantly higher than those receiving the shorter (19-29 days) BNT162b2 schedule (694; 95%CI: 540-893). Unlike BNT162b2 recipients, there was no decline in antibody titres among AZD1222 recipients between 7-13 and 14-34 days post-dose 2 regardless of interval. Responses were two-fold higher among AZD1222 recipients with a 65-84 compared with 45-64 day dose interval. Responses were, however, lower following an 85+ day interval between AZD1222 doses, although this group was small, with older participants and lower dose 1 responses.

**Table 4:** Adjusted models at two time points post dose 2, using AZ 65-84 day schedule group as reference

|  | 7-13 days post dose 2 |  | 14-34 days post dose 2 |  |
| --- | --- | --- | --- | --- |
|  | geometric mean<br>ratio of (log)<br>RocheS responses | p-value | geometric mean<br>ratio of (log)<br>RocheS responses | p-value |
| vaccine & schedule |  |  |  |  |
| Astra Zeneca, 45-64 days | 0.64 (0.4 - 1.04) | 0.069 | 0.51 (0.34 - 0.77) | 0.001 |
| Astra Zeneca, 65-84 days | 1 (ref) |  | 1 (ref) |  |
| Astra Zeneca, 85+ days | - |  | 0.62 (0.33 - 1.18) | 0.148 |
| Pfizer, 19-29 days | - |  | 0.66 (0.45 - 0.97) | 0.036 |
| Pfizer, 65-84 days | 8.29 (5.39 - 12.74) | <0.001 | 6.38 (4.56 - 8.92) | <0.001 |
| Pfizer, 85+ days | 17.21 (7.16 - 41.38) | <0.001 | - |  |
| age group |  |  |  |  |
| 50-64 | 1 (ref) |  | 1 (ref) |  |
| 65-79 | 1.13 (0.68 - 1.86) | 0.634 | 0.83 (0.48 - 1.43) | 0.438 |
| 80-89 | 0.95 (0.48 - 1.86) | 0.877 | 1.16 (0.51 - 2.62) | 0.683 |
| sex |  |  |  |  |
| male | 1 (ref) |  | 1 (ref) |  |
| female | 1.68 (1.24 - 2.28) | 0.001 | 1.08 (0.79 - 1.47) | 0.019 |

In all groups, GMTs after two vaccine doses regardless of interval were higher than those observed after mild-to-moderate COVID-19 (Table 3, Figure 3).

In participants previously infected, following dose 2 antibodies were further boosted by BNT162b2, increasing to 27322.5 (95%CI: 17444.4 - 42794.2), but not by AZD1222, where the GMT was 9633.2 (95%CI: 6233.9-14886.3) at 14-34 days post-dose 2.

### Post dose 2: Vaccine effectiveness following extended schedules

VE was higher across all age-groups for both vaccines from 14 days after dose 2 compared to dose 1 but the magnitude depended on the interval between doses (Figure 4). Amongst BNT162b2 recipients, VE was consistently higher with >45 day intervals compared to 19-29 days for all age-groups.

**Figure 4:**
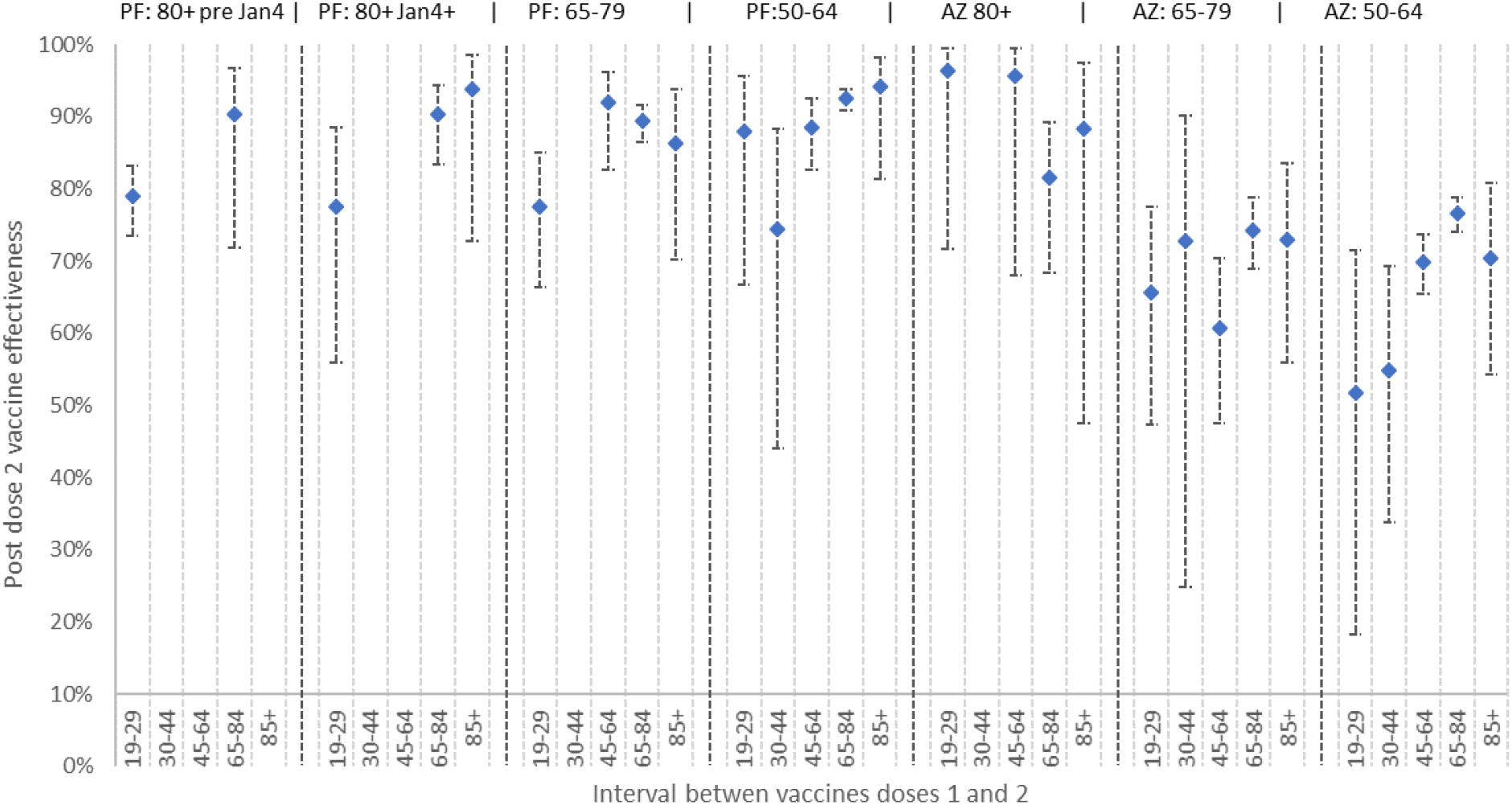
Two dose vaccine effectiveness by age group, vaccine type and interval between doses.

Amongst AZD1222 recipients aged ≥80 years, two-dose VE after 14 days was 96% (68-99) and 82% (95%CI: 68-89) following 45-64 and 65-84 days intervals, respectively (Figure 4). Those receiving their second dose outside of these recommended intervals also had high VE after two doses; for ≥85 day interval, the estimated VE was 88% (95%CI: 48-97). In younger adults, two-dose VE was higher but not statistically significant with a 65-84 than a 45-64 day interval, but lower at all timepoints than ≥80 year-olds (Figure 4, Table Supplementary files).

## Discussion

Our findings uniquely combine serological and vaccine effectiveness data for extended immunisation schedules in adults who were prioritised for vaccination during the first phase of the UK COVID-19 immunisation programme. We demonstrate high and sustained antibodies responses for >12 weeks after the first dose of either BNT162b2 or AZD1222 vaccine in previously-uninfected adults, with 97.7% and 95.6%, respectively, becoming seropositive by 35-55 days after their first dose. Antibody levels rose more rapidly and then stabilised after a single BNT162b2 dose but increase more gradually after AZD1222, such that antibody levels were equivalent in both cohorts by 56-76 days after a single dose. In previously-infected individuals, both vaccines provided significant boosting after one dose, with S-antibody GMTs >50 fold higher than adults with mild-to-moderate COVID at 8-12 weeks post-infection. These serological findings are consistent with national surveillance data on clinical protection against symptomatic disease. VE after a single BNT162b2 dose was 53-58% after 28 days across all age-groups, with no evidence of a decline in effectiveness with age and only a modest decline in effectiveness beyond 56-76 days after the first dose. For AZD1222, single-dose VE was 43-55% beyond 28 days, with some evidence of a decline in amongst the oldest age-group beyond 10 weeks.

The UK decision to recommend extended COVID-19 vaccine schedules against emergency use authorisation was considered highly controversial in the midst of a large second wave caused by the more transmissible Alpha variant. Lengthening intervals between vaccine doses to enhance boosting is well-recognised and was demonstrated in the pre-licensure AZD1222 trial,(13, 14) but the lack of similar data for extended schedules using BNT162b2 prompted a rapid post-implementation evaluation to monitor serological responses and real world effectiveness data for both COVID-19 vaccines in the UK.

With BNT162b2, we found 8-10 fold higher GMTs after the second dose with a 6-9 or 10-13 week interval compared with the authorised 3-week interval, which was also associated with more rapid waning of up to 50% between 1.5-3 weeks and 19 weeks after dose 2. These findings are consistent with our other as-yet unpublished study in ≥80 year-olds, where peak antibody responses were 3.5-fold higher following extended-schedule BNT162b2 immunisation although, interestingly, cellular responses were 3.6-fold lower with the extended interval schedule.(15) The current study, which includes a wider age-range, found no evidence of declining antibody with age after either dose, although a recent study reported lower serum neutralisation and binding IgG/IgA after the first dose with increasing age, and lower potency against SARS-CoV-2 variants of concern (VOC) among ≥80 year-olds.(16) Following the second dose, neutralisation against VOC was detectable regardless of age.

Among AZD1222, the initial clinical trials allowed permissive interval between vaccine doses which showed minimal waning of antibodies or protection against symptomatic COVID-19 for up to 3 months after the first dose in healthy working-age adults, with better boosting after the second dose after a longer interval.(17) In our cohort, too, which focused on older adults, AZD1222 recipients had 2-fold higher antibodies after a 10-13 compared to 6-9 week interval.

In previously-infected adults, we observed significant boosting of antibody responses after the first dose of both vaccines and after the second dose of BNT162b2, but not AZD1222. Notably, though, antibody GMTs following two doses of either vaccine were higher than those observed following mild-to-moderate COVID-19 regardless of interval.

We were also able to compare immunogenicity with real-world VE data in England, which show substantial protection against symptomatic disease from 14 days after dose 2. Higher two-dose VE was observed with >6 week intervals between BNT162b2 doses compared to the authorised 3-week schedule, including ≥80 year-olds. Among AZD1222 recipients, two doses provided the highest protection among ≥80 year-olds regardless of interval. Surprisingly, two-dose VE among 50-64 year-olds was lower, even when compared with adults in the same-age group receiving two BNT162b2 doses. This may be due to the differential use of the vaccines in the national immunisation programme-because AZD1222 vaccines do not require ultra-low temperatures for storage or transport, they were preferred for vaccinating care home residents who a higher risk of natural exposure and pre-existing immunity prior to vaccination, and for clinical risk groups in the community.(18) At the same time, BNT162b2 was preferentially used for healthy healthcare workers in hospitals early in the national vaccine rollout. Since BNT162b2 was deployed earlier than AZD1222, we have longer population follow-up for this vaccine. Whilst the analyses adjust for key confounding variables, such as period and risk groups, whilst excluding those with previous COVID, it is possible that residual confounding persists to some extent. Additionally, the Alpha variant was dominant during the majority of the study period. Since May 2021, this has been replaced by the Delta variant, and differences in VE between Alpha and Delta have been reported. (19, 20)

Notwithstanding this, it is important to emphasise that a single dose of either vaccine remains highly effective against severe endpoints, which is the primary aim of the vaccination programme, with 75-85% protection against hospitalisation in the oldest cohorts.(8) As of 28/06/2021, the vaccination programme is estimated to have prevented nearly 8 million infections and 27,000 deaths in England alone.(21, 22)

### Strengths and Limitations

The strength of this study is the combination of sero-surveillance with real-world national VE data for two different vaccines in different age groups, including older adults who were excluded from initial clinical trials, with variable, real-world dosing intervals. Serological assessments provide an objective measure of vaccine responses which are important for comparing vaccines and schedules, but interpretation of serological data is limited as the way in which it correlates with protection is unknown and the recognition that neutralising activity of antibodies and cellular immunity also play an important role in protection. As with any observational study, there are limitations to VE analysis. There may be confounding factors that could increase the risk of COVID-19 in vaccinated individuals, for example, if vaccinated individuals adopted riskier behaviours after vaccination or unvaccinated individuals isolated themselves to reduce their risk of viral exposures. In addition, VE could be attenuated if there are high levels of protection from previous infection in the population or if there is misclassification of cases and test negative controls due to low sensitivity or specificity of PCR-testing.

### Conclusions

Our findings support the UK decision to prioritise the first dose by delaying the second dose of COVID-19 vaccine to maximise public health impact. Given the global vaccine constraints, these results are relevant to policymakers in low and middle income countries especially in the context of highly transmissible variants and rising incidence in many parts of the world. An additional yet undervalued benefit of extended schedules is higher boosting and better protection after two doses of either vaccine, which potentially confer better protection against variants and for a longer duration than short-interval schedules. Our data also confirm previous findings of high protection after a single vaccine dose in previously-infected individuals, which is also important in the context of limited vaccine supplies. Ongoing evaluation of the protection conferred against new variants using an extended schedule will be critical.

## Data Availability

Applications for relevant anonymised data should be submitted to the Public Health England Office for Data Release.

https://www.gov.uk/government/publications/accessing-public-health-england-data/about-the-phe-odr-and-accessing-data

## Acknowledgments

We would like to thank Dorothy Blundell, Dr Caroline Sayer and the team at Haverstock Healthcare GP Federation, and the whole CONSESUS team at PHE including those within the Virus Reference Department at Colindale who assisted with the laboratory testing.

## Funding

This surveillance was funded by Public Health England. The CONSENSUS study/audit was approved by PHE’s R&D Research Ethics and Governance Group. No: NR0253

## Author Contributions

SL, FB, PM, MR, KB, GA conceived and designed the study; LW and EL supervised the laboratory testing; SS, CW, MO’B and FB co-ordinated the patient enrolment. NJA and HW performed the statistical analysis and GA, JLB, SL, and KB wrote the manuscript. All authors read and approved the submission.

## Declaration of Interests

MER reports that the Immunisation and Countermeasures Division (PHE) has provided vaccine manufacturers with post-marketing surveillance reports on pneumococcal and meningococcal infection, which the companies are required to submit to the UK licensing authority in compliance with their risk management strategy. A cost-recovery charge is made for these reports. EL report that the PHE Vaccine Evaluation Unit does contract research on behalf of GlaxoSmithKline, Sanofi, and Pfizer, which is outside the submitted work.

## Notes

### Author Declarations

This surveillance was funded by Public Health England. The CONSENSUS study/audit was approved by PHEs R&D Research Ethics and Governance Group. No NR0253

