## Supplementary Materials for "Higher serological responses and increased vaccine effectiveness demonstrate the value of extended vaccine schedules in combatting COVID-19 in England"

**Supplementary Material**

**Supplementary Table 1:** Demographics of individuals in the CONSENSUS study, by vaccine and age group.

| Vaccine & age group | contribution | N | age | sex (if known) | ethnicity (if known) | past infection |
| --- | --- | --- | --- | --- | --- | --- |
|  |  |  | median (IQR) | n male (%) | n non-white (%) | n (%) |
| AstraZeneca, extended schedule, ages 50-64 | all recruited | 170 | 54 (52 - 57) | 81 (48.8%) | 89 (52.4%) | 56 (32.9%) |
|  | ≥2 dose 1 samples | 111 | 55 (53 - 58) | 51 (46.4%) | 48 (43.2%) | 36 (32.4%) |
|  | ≥1 dose 2 sample | 80 | 54 (53 - 57.5) | 39 (49.4%) | 26 (32.5%) | 23 (28.8%) |
| AstraZeneca, extended schedule, ages 65-79 | all recruited | 147 | 70 (68 - 73) | 68 (46.3%) | 36 (24.7%) | 25 (17%) |
|  | ≥2 dose 1 samples | 121 | 70 (68 - 73) | 54 (44.6%) | 26 (21.7%) | 20 (16.5%) |
|  | ≥1 dose 2 sample | 111 | 70 (68 - 73) | 47 (42.3%) | 19 (17.3%) | 16 (14.4%) |
| Astra Zeneca, extended schedule, ages 80-89 | all recruited | 12 | 84 (82 - 85.5) | 5 (41.7%) | 2 (16.7%) | 1 (8.3%) |
|  | ≥2 dose 1 samples | 10 | 83 (82 - 85) | 3 (30%) | 1 (10%) | 1 (10%) |
|  | ≥1 dose 2 sample | 9 | 82 (82 - 84) | 3 (33.3%) | 1 (11.1%) | 1 (11.1%) |
| Pfizer, extended schedule, ages 50-64 | all recruited | 1 | 61 | 0 |  | 0 |
|  | ≥2 dose 1 samples | 1 | 61 | 0 |  | 0 |
|  | ≥1 dose 2 sample | 1 | 61 | 0 |  | 0 |
| Pfizer, extended schedule, ages 65-79 | all recruited | 260 | 72 (69 - 74) | 117 (45%) | 28 (10.9%) | 26 (10%) |
|  | ≥2 dose 1 samples | 211 | 71 (69 - 74) | 95 (45%) | 19 (9.1%) | 19 (9%) |
|  | ≥1 dose 2 sample | 225 | 72 (69 - 74) | 102 (45.3%) | 22 (9.9%) | 21 (9.3%) |
| Pfizer, extended schedule, ages 80-89 | all recruited | 72 | 82 (81 - 84) | 31 (43.1%) | 7 (9.9%) | 9 (12.5%) |
|  | ≥2 dose 1 samples | 63 | 82 (81 - 84) | 25 (39.7%) | 6 (9.7%) | 7 (11.1%) |
|  | ≥1 dose 2 sample | 56 | 83 (81 - 84) | 22 (39.3%) | 4 (7.3%) | 5 (8.9%) |
| Pfizer, standard schedule, ages 65-79 | all recruited | 59 | 75 (74 - 77) | 28 (47.5%) | 6 (10.3%) | 7 (11.9%) |
|  | ≥2 dose 1 samples | 1 | 66 | 0 | 0 | 0 |
|  | ≥1 dose 2 sample | 59 | 75 (74 - 77) | 28 (47.5%) | 6 (10.3%) | 7 (11.9%) |
| Pfizer, standard schedule, ages 80-89 | all recruited | 29 | 82 (81 - 85) | 14 (48.3%) | 3 (10.3%) | 1 (3.4%) |
|  | ≥2 dose 1 samples | 0 |  |  |  |  |
|  | ≥1 dose 2 sample | 28 | 82 (80.5 - 85) | 14 (50%) | 3 (10.7%) | 0 (0%) |

**Supplementary Table 2: Test negative case control results showing adjusted odds ratios (OR) and vaccine effectiveness (VE) post first dose and post second dose according to intervals between doses for Pfizer vaccine in different age cohorts.**

| Pfizer  dose | days between doses | days since dose | controls | cases | OR (95% CI) | VE (95% CI) |
| --- | --- | --- | --- | --- | --- | --- |
| Age 80+ and first vaccine dose before January 4th 2021 or unvaccinated. Cases and controls from Dec 8th 2020 | | | | | | |
|  |  | unvaccinated | 15936 | 9178 | base | base |
| 1 |  | 0-3 | 281 | 172 | 1.23 (1.01-1.5) |  |
|  |  | 4-13 | 862 | 731 | 1.29 (1.16-1.44) |  |
|  |  | 14-27 | 754 | 467 | 0.9 (0.8-1.02) | 10% (-2-20) |
|  |  | >=28 | 1528 | 269 | 0.45 (0.38-0.53) | 55% (47-62) |
| 2 | 19-29 | >=14 | 2219 | 144 | 0.21 (0.17-0.26) | 79% (74-83) |
|  | 30-44 | >=14 | 6 | 0 |  |  |
|  | 45-64 | >=14 | 22 | 0 |  |  |
|  | 65-84 | >=14 | 522 | 4 | 0.1 (0.03-0.28) | 90% (72-97) |
|  | 85+ | >=14 | 13 | 0 |  |  |
| Age 80+ and first vaccinated from Jan 4th 2021 or unvaccinated, cases and controls from Jan 4th 2021 | | | | | | |
|  |  | unvaccinated | 5676 | 3881 | base | base |
| 1 |  | 0-3 | 424 | 267 | 0.98 (0.83-1.15) |  |
|  |  | 4-13 | 1277 | 699 | 0.93 (0.83-1.05) |  |
|  |  | 14-27 | 1580 | 398 | 0.59 (0.51-0.68) | 41% (32-49) |
|  |  | >=28 | 3315 | 385 | 0.58 (0.48-0.69) | 42% (31-52) |
| 2 | 19-29 | >=14 | 212 | 22 | 0.23 (0.12-0.44) | 77% (56-88) |
|  | 30-44 | >=14 | 5 | 0 |  |  |
|  | 45-64 | >=14 | 42 | 1 | 0.18 (0.02-1.34) | 82% (-34-98) |
|  | 65-84 | >=14 | 1289 | 30 | 0.1 (0.06-0.17) | 90% (83-94) |
|  | 85+ | >=14 | 83 | 2 | 0.06 (0.01-0.27) | 94% (73-99) |
| Age 65-79 and first vaccinated from Jan 4th 2021 or unvaccinated, cases and controls from Jan 4th 2021 | | | | | | |
|  |  | unvaccinated | 76821 | 26463 | base | base |
| 1 |  | 0-3 | 1536 | 600 | 1.3 (1.18-1.43) |  |
|  |  | 4-13 | 4338 | 1242 | 1.06 (0.99-1.14) |  |
|  |  | 14-27 | 4896 | 640 | 0.57 (0.52-0.63) | 43% (37-48) |
|  |  | >=28 | 11053 | 745 | 0.47 (0.42-0.52) | 53% (48-58) |
| 2 | 19-29 | >=14 | 388 | 33 | 0.23 (0.15-0.34) | 77% (66-85) |
|  | 30-44 | >=14 | 49 | 0 |  |  |
|  | 45-64 | >=14 | 483 | 7 | 0.08 (0.04-0.17) | 92% (83-96) |
|  | 65-84 | >=14 | 5247 | 118 | 0.11 (0.08-0.14) | 89% (86-92) |
|  | 85+ | >=14 | 174 | 7 | 0.14 (0.06-0.3) | 86% (70-94) |
| Age 50-64 and first vaccinated from Feb 1st 2021 or unvaccinated, cases and controls from Feb 1st 2021 | | | | | | |
|  |  | unvaccinated | 136905 | 33904 | base | base |
| 1 |  | 0-3 | 1478 | 377 | 1.09 (0.97-1.22) |  |
|  |  | 4-13 | 3599 | 831 | 1.04 (0.96-1.12) |  |
|  |  | 14-27 | 4116 | 457 | 0.53 (0.48-0.59) | 47% (41-52) |
|  |  | >=28 | 9300 | 702 | 0.49 (0.45-0.53) | 51% (47-55) |
| 2 | 19-29 | >=14 | 83 | 4 | 0.12 (0.04-0.33) | 88% (67-96) |
|  | 30-44 | >=14 | 114 | 7 | 0.26 (0.12-0.56) | 74% (44-88) |
|  | 45-64 | >=14 | 643 | 24 | 0.11 (0.08-0.17) | 89% (83-92) |
|  | 65-84 | >=14 | 3977 | 127 | 0.08 (0.06-0.09) | 92% (91-94) |
|  | 85+ | >=14 | 110 | 3 | 0.06 (0.02-0.19) | 94% (81-98) |

**Supplementary Table 3: Test negative case control results showing adjusted odds ratios (OR) and vaccine effectiveness (VE) post first dose and post second dose according to intervals between doses for Astra Zeneca vaccine in different age cohorts.**

| AZ dose | days between doses | days since dose | controls | cases | OR (95% CI) | VE (95% CI) |
| --- | --- | --- | --- | --- | --- | --- |
| Age 80+ and first vaccinated from Jan 4th 2021 or unvaccinated, cases and controls from Jan 4th 2021 | | | | | | |
|  |  | unvaccinated | 5676 | 3881 | base | base |
| 1 |  | 0-3 | 424 | 228 | 0.74 (0.62-0.88) |  |
|  |  | 4-13 | 853 | 545 | 1.03 (0.9-1.19) |  |
|  |  | 14-27 | 957 | 333 | 0.78 (0.66-0.93) | 22% (7-34) |
|  |  | >=28 | 1986 | 237 | 0.58 (0.47-0.71) | 42% (29-53) |
| 2 | 19-29 | >=14 | 53 | 1 | 0.04 (0-0.28) | 96% (72-100) |
|  | 30-44 | >=14 | 5 | 1 |  |  |
|  | 45-64 | >=14 | 92 | 1 | 0.04 (0.01-0.32) | 96% (68-99) |
|  | 65-84 | >=14 | 574 | 31 | 0.18 (0.11-0.32) | 82% (68-89) |
|  | 85+ | >=14 | 39 | 2 | 0.12 (0.03-0.52) | 88% (48-97) |
| Age 65-79 and first vaccinated from Jan 4th 2021 or unvaccinated, cases and controls from Jan 4th 2021 | | | | | | |
|  |  | unvaccinated | 76821 | 26463 | base | base |
| 1 |  | 0-3 | 3692 | 636 | 0.68 (0.62-0.75) |  |
|  |  | 4-13 | 6261 | 1631 | 1.1 (1.03-1.17) |  |
|  |  | 14-27 | 7070 | 937 | 0.67 (0.61-0.73) | 33% (27-39) |
|  |  | >=28 | 17000 | 1045 | 0.48 (0.44-0.54) | 52% (46-56) |
| 2 | 19-29 | >=14 | 247 | 30 | 0.34 (0.23-0.53) | 66% (47-77) |
|  | 30-44 | >=14 | 101 | 4 | 0.27 (0.1-0.75) | 73% (25-90) |
|  | 45-64 | >=14 | 945 | 68 | 0.39 (0.3-0.53) | 61% (47-70) |
|  | 65-84 | >=14 | 7556 | 435 | 0.26 (0.21-0.31) | 74% (69-79) |
|  | 85+ | >=14 | 270 | 20 | 0.27 (0.17-0.44) | 73% (56-83) |
| Age 50-64 and first vaccinated from Feb 1st 2021 or unvaccinated, cases and controls from Feb 1st 2021 | | | | | | |
|  |  | unvaccinated | 136905 | 33904 | base | base |
| 1 |  | 0-3 | 6895 | 1012 | 0.7 (0.65-0.75) |  |
|  |  | 4-13 | 12120 | 2584 | 1.1 (1.05-1.16) |  |
|  |  | 14-27 | 14597 | 1491 | 0.62 (0.58-0.66) | 38% (34-42) |
|  |  | >=28 | 34222 | 3633 | 0.58 (0.54-0.61) | 42% (39-46) |
| 2 | 19-29 | >=14 | 84 | 18 | 0.48 (0.29-0.82) | 52% (18-71) |
|  | 30-44 | >=14 | 219 | 33 | 0.45 (0.31-0.66) | 55% (34-69) |
|  | 45-64 | >=14 | 2311 | 313 | 0.3 (0.26-0.34) | 70% (66-74) |
|  | 65-84 | >=14 | 7443 | 782 | 0.23 (0.21-0.26) | 77% (74-79) |
|  | 85+ | >=14 | 165 | 25 | 0.3 (0.19-0.46) | 70% (54-81) |
